# Medicinal Cannabis Plant Extract (NTI164) modifies epigenetic, ribosomal, and immune pathways in paediatric acute-onset neuropsychiatric syndrome

**DOI:** 10.1101/2025.06.30.25330605

**Authors:** Brooke A Keating, Velda X Han, Hiroya Nishida, Nader Aryamanesh, Lee L Marshall, Brian S Gloss, Xianzhong Lau, Ruwani Dissanayake, Suat Dervish, Mark E Graham, Shekeeb S Mohammad, Manoj Kanhangad, Michael C Fahey, Shrujna Patel, Russell C Dale

## Abstract

**Background:** Paediatric acute-onset neuropsychiatric syndrome (PANS) is a syndrome of infection-provoked abrupt-onset obsessive-compulsive disorder (OCD) or eating restriction. Based on the hypothesis that PANS is an epigenetic disorder of immune and brain function, a full-spectrum medicinal cannabinoid-rich low-THC cannabis (NTI164) was selected for its known epigenetic and immunomodulatory properties.

**Methods:** This open-label trial of fourteen children with chronic-relapsing PANS (mean age 12.1 years; range 4–17; 71% male) investigated the safety and efficacy of 20mg/kg/day NTI164 over 12 weeks. Clinical outcomes were assessed using gold standard tools. To define the biological effects of NTI164, blood samples were collected pre- and post-treatment for bulk and single-cell transcriptomics, proteomics, phosphoproteomics, and DNA methylation.

**Findings:** NTI164 was well-tolerated, and 12 weeks of treatment decreased the mean Clinical Global Impression-Severity (CGI-S) score from 4.8 to 3.3 (p=0.002). Significant improvements were observed in emotional regulation (RCADS-P, p<0.0001), obsessive-compulsive disorder (CYBOCS-II, p=0.0001), tics (YGTSS, p<0.0001), attention-deficit hyperactivity disorder (Conner’s, p=0.028), and overall quality of life (EQ-5D-Y, p=0.011). At baseline, the multi-omic approach revealed that leukocytes from patients with PANS had dysregulated epigenetic (chromatin structure, DNA methylation, histone modifications, transcription factors), ribosomal, mRNA processing, immune, and signalling pathways. These pathways were significantly modulated by NTI164 treatment.

**Interpretation:** NTI164 shows promise as a disease-modifying therapeutic for PANS. Multi-omics reveal broad epigenetic and immune dysregulation in patients, which was modified by NTI164, presenting epigenetic machinery as a therapeutic target in PANS.

**Funding:** This study was funded by Fenix Innovation Group Pty Ltd and Neurotech International Ltd.

## INTRODUCTION

Paediatric acute-onset neuropsychiatric syndrome (PANS) is a debilitating condition, characterised by abrupt-onset obsessive-compulsive disorder (OCD) or severely restrictive food intake with co-existing symptoms, including anxiety, depression, irritability, behavioural regression, cognitive deterioration, sensory or motor abnormalities, and somatic symptoms (1). PANS often has a relapsing-remitting clinical course where periods of stability can be followed by relapses (2), triggered by infections or other stressors.

A recent American Academy of Pediatrics statement recognises PANS as a clinical syndrome, however it is unclear whether PANS is a distinct neuroimmune entity or part of the neurodevelopmental continuum (3, 4). OCD affects 2% of children and commonly co-exists with other neurodevelopmental disorders (NDDs), such as autism spectrum disorder, attention-deficit hyperactivity disorder (ADHD), and Tourette syndrome (5). Rare, pathogenic DNA variants are found in only a small minority of patients with NDDs (6). Instead, a combination of genetic and environmental factors is considered to contribute to the development of NDDs, through epigenetic, gene regulatory, and immune processes (7, 8). We propose that PANS is a clinical phenotype driven by gene-environment interactions, involving epigenetic dysregulation that impacts both the immune system and brain (9, 10). There are currently no approved treatments for PANS, and patients often have refractory symptoms, despite the use of conventional psychotherapy or psychiatric medications.

*Cannabis sativa L.* has long been used in medicine, and increasingly proposed as treatment of psychiatric disorders and NDDs (11, 12). The plant secretes a resin which contains a mix of cannabinoids with two principal components, Δ9-tetrahydrocannabinol (THC) and cannabidiol (CBD). Extensive research centres around CBD, which has generated much interest due to a lack of psychoactive activity (unlike THC) and excellent tolerability in humans, as well as anti-inflammatory capabilities (13). While THC can elicit undesired psychoactive effects, emerging evidence suggests cannabis extracts with all components of the plant are more potent immunomodulators than CBD alone (or other isolated compounds), termed the “entourage effect” (14). Other studies support the observation that botanical drugs are more efficacious than their isolated components (15).

In this open-label study (FENPANS1, ClinicalTrials.gov ID: NCT06621888), the safety and efficacy of NTI164, a full-spectrum medicinal cannabis plant derived from proprietary strains of *Cannabis sativa* and containing cannabidiolic acid (CBDA), CBD, cannabigerolic acid (CBGA), cannabidivarin (CBDV), and extremely low THC (0.08%) were investigated in a cohort of children diagnosed with PANS. NTI164 was administered orally twice daily at a dose of 20mg/kg/day for 12 weeks. This open-label study measured changes in PANS clinical symptoms and explored the biological effects of NTI164 on peripheral immune cells through cytokine assays, transcriptomic, proteomic, phosphoproteomic, and DNA methylation analyses, at baseline and after treatment with NTI164 in children with PANS.

## RESULTS

### Clinical characteristics of children with PANS

#### Family history

A total of 14 patients fulfilling PANS criteria (1, 16) (full inclusion/exclusion criteria available in Appendix 1) from New South Wales and Victoria, Australia were recruited to the study from two tertiary neurology clinics. The mean age was 12.1 (range 4-17) years ((4 females, 29%; 10 males, 71%), Table 1), 64.3% (n = 9) of the probands had a first-degree relative with an autoimmune condition, and 50% (n = 7) of probands had a first-degree relative with a neurodevelopmental or neuropsychiatric disorder (Table 1).

#### PANS symptoms

The mean age of PANS onset was 5.2 years (range 1.5-12 years) and was typically triggered by infections (n = 12, 85.7%). In our cohort, all patients demonstrated a relapsing-remitting phenotype and had an average of five PANS-related flares each year (range 2-8 flares a year), lasting 3-4 days to 6 months in duration, which were mainly infection-provoked. The PANS symptoms in our cohort were OCD (n = 14, 100%), anxiety (n = 8, 57.1%), regression/cognitive decline (n = 8, 57.1%), inattention or agitation (n = 6, 42.9%), tics/Tourette syndrome (n = 3, 21.4%), rage or oppositional behaviours (n = 3, 21.4%), and severe food restriction (n = 3, 21.4%). In our cohort, other NDD and neuropsychiatric diagnoses diagnosed, in addition to their PANS diagnosis, included autism spectrum disorder (n = 8, 53.3%) and ADHD (n = 6, 42.9%). The average duration between PANS onset and enrolment into the current study was 6.7 (range 2-13) years.

#### Severity of PANS symptoms

At baseline, the mean Clinical Global Impression – Severity (CGI-S) score was 4.8 out of 7 (markedly ill; score range 4-6, higher score indicates severe disease), and baseline Revised Children’s Anxiety and Depression Scale – Parent-rated (RCADS-P) and Children’s Yale-Brown Obsessive-compulsive Scale (CY-BOCS) scores were 101.9 (score range 92-129) and 33.1 (score range 24-48) respectively, indicating severe emotional disorders and OCD.

#### Neuropsychiatric and PANS treatments

All patients had previously received or were receiving ongoing psychological support. Children were taking an average of 2.4 medications (range 1-5) at the time of trial enrolment, including selective serotonin reuptake inhibitors (SSRIs; n = 9, 64.3%), antipsychotics (n = 9, 64.3%), stimulants (n = 7, 50%), and α-agonists (n = 3, 21.4%). All patients had previously received antibiotics as a rescue treatment for acute episodes. Additionally, 14.3% (n=2) had been treated with steroids or intravenous immunoglobulin (IVIg) during prior flares. As part of inclusion criteria to the current study, IVIg, steroids, and antibiotic use was not permitted for at least 12 weeks before entering the study or throughout the study period.

### Safety and tolerability of NTI164 in PANS patients

Oral NTI164 was well-tolerated in patients at 20mg/kg/day, divided into 2 doses, with no significant abnormalities observed in laboratory results (full blood examination, including kidney and liver function tests, Supplementary Table 1) after 12 weeks of treatment. Adverse events (AEs) determined to be related to the Investigational Product (Supplementary Table 2) were experienced by 33% of patients, with these being symptoms of gastrointestinal symptoms (i.e. nausea, diarrhoea), lethargy, or changes to sleep behaviour (i.e. taking longer to fall asleep). One patient required dose-adjustment due to diarrhoea (reduced to 15mg/kg/day) and one patient due to lethargy (reduced to 17.5mg/kg/day). All other AEs were self-resolving and did not impact the patients’ functioning. No patients discontinued NTI164 due to AEs during the study period.

### NTI64 improved clinical symptoms associated with PANS

Several gold-standard assessments of behaviour and tics (Supplementary Table 3) were implemented in the FENPANS1 study and were assessed by trained personnel at baseline and following 12 weeks of NTI164 oral administration (Figure 1, Supplementary Table 4). At baseline, the mean CGI-S score was 4.8 (range 4-6), and was significantly reduced (i.e. improved) to 3.3 (range 2-5) after 12 weeks of NTI164 administration (Figure 1A, p = 0.002). Similarly, NTI164 administration significantly improved emotional regulation (RCADS-P, Figure 1B, p < 0.0001), OCD (CYBOCS-II, Figure 1C, p = 0.0001), tics (YGTSS, Figure 1D, p < 0.0001), ADHD (Conner’s, Figure 1E, p = 0.028), and overall quality of life (EQ-5D-Y, Figure 1F, p = 0.011). Notably, the RCADS-P showed improvements in all subdomains, including social phobia (p < 0.0001), panic disorder (p = 0.018), major depression (p = 0.0098), separation anxiety (p < 0.0001), generalised anxiety (p < 0.0001), and obsessive-compulsive symptoms (p = 0.001) (subdomains for the relevant surveys can be found in Supplementary Figure 1 and Supplementary Table 4). Following 12 weeks of NTI164 treatment, 13 patients/caregivers (85.7%) elected to continue receiving NTI164 into an Extension phase.

**Figure 1.**
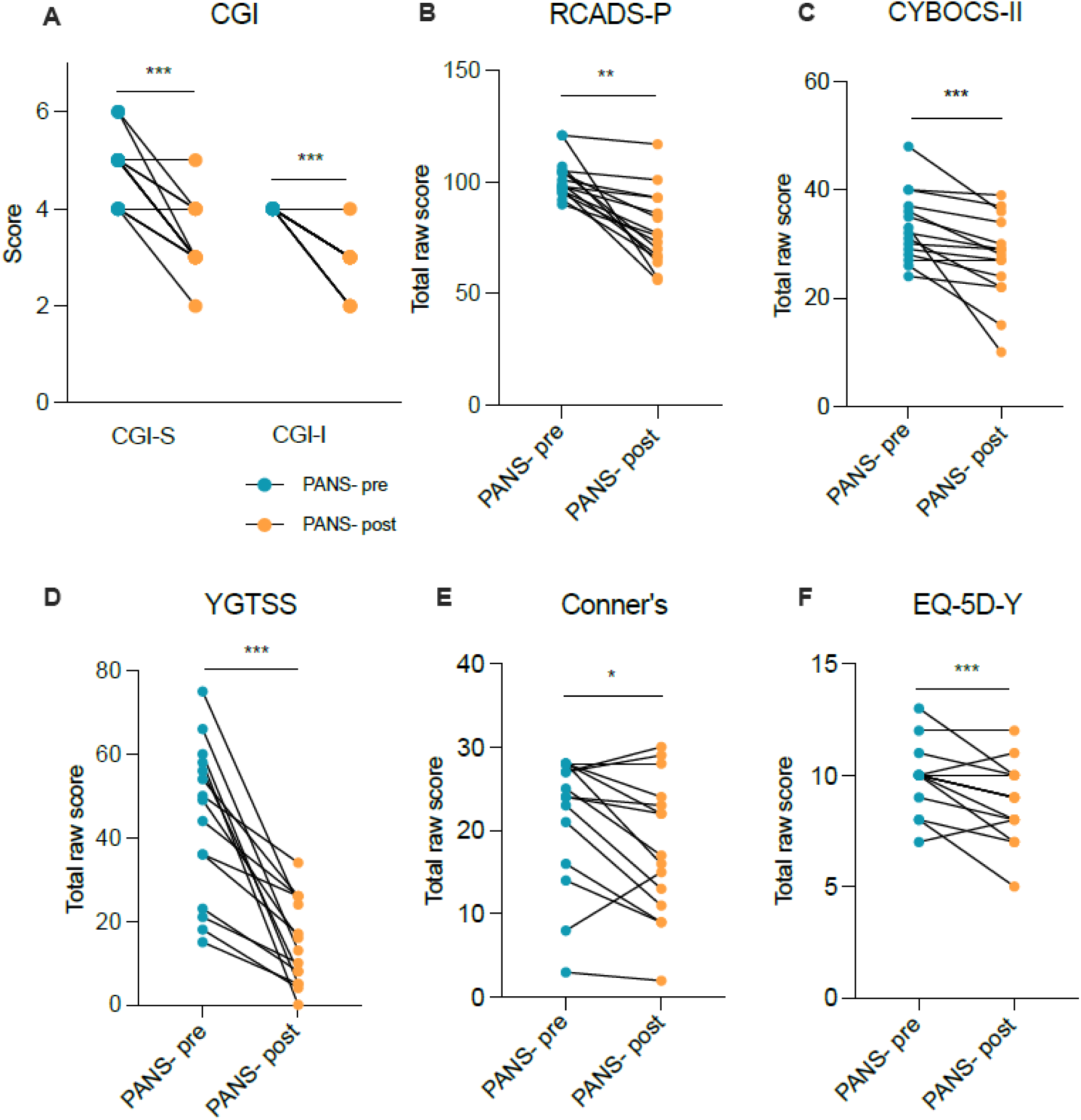
Clinical survey scores of PANS children at baseline (PANS-pre) were significantly improved following 12 weeks of NTI164 treatment (PANS-post). (A) Clinical Global Impression - Severity (CGI-S) and Clinical Global Impression - Improvement (CGI-I), CGI-S mean score PANS-pre = 4.8, mean score PANS-post = 3.3, mean CGI-I score = 2.8 between PANS-pre and PANS-post, n = 14, some data points represent multiple patients (e.g. several patients scoring 5 at baseline and scoring 4 post-NTI164). (B) Revised Children’s Anxiety and Depression Scale - Parent-rated (RCADS-P), mean score PANS-pre = 101.9, mean score PANS-post = 78.6. (C) Children’s Yale-Brown Obsessive Compulsive Scale - 2nd version (CYBOCS-II), mean score PANS- pre = 33.1, mean score PANS-post = 27.3. (D) Yale Global Tic Severity Scale (YGTSS), mean score PANS-pre = 44.1, mean score PANS-post = 14.8. (E) Conner’s scale, mean score PANS-pre = 21.6, mean score PANS-post = 18. (F) EQ-5D-Y, mean score PANS-pre = 9.9, mean score PANS-post = 8.8 Wilcoxon matched-pairs signed rank test, n = 14, *P < 0.05, **P < 0.01, ***P < 0.001, ****P < 0.0001.

### PANS children have dysregulated immune cell transcriptomic signatures, which were modified by NTI164

#### Cytokine assay

A multiplex 13-cytokine assay performed on plasma revealed elevated brain-derived neurotrophic factor (BDNF) (Figure 2A, p = 0.0014) and interleukin-(IL)-18 (Figure 2B, p = 0.0266) at baseline (PANS-pre) compared to controls. Other cytokines in the assay were generally increased in PANS-pre and reduced in patients with PANS post-NTI164, but no additional significant differences were found (Supplementary Figure 2).

**Figure 2.**
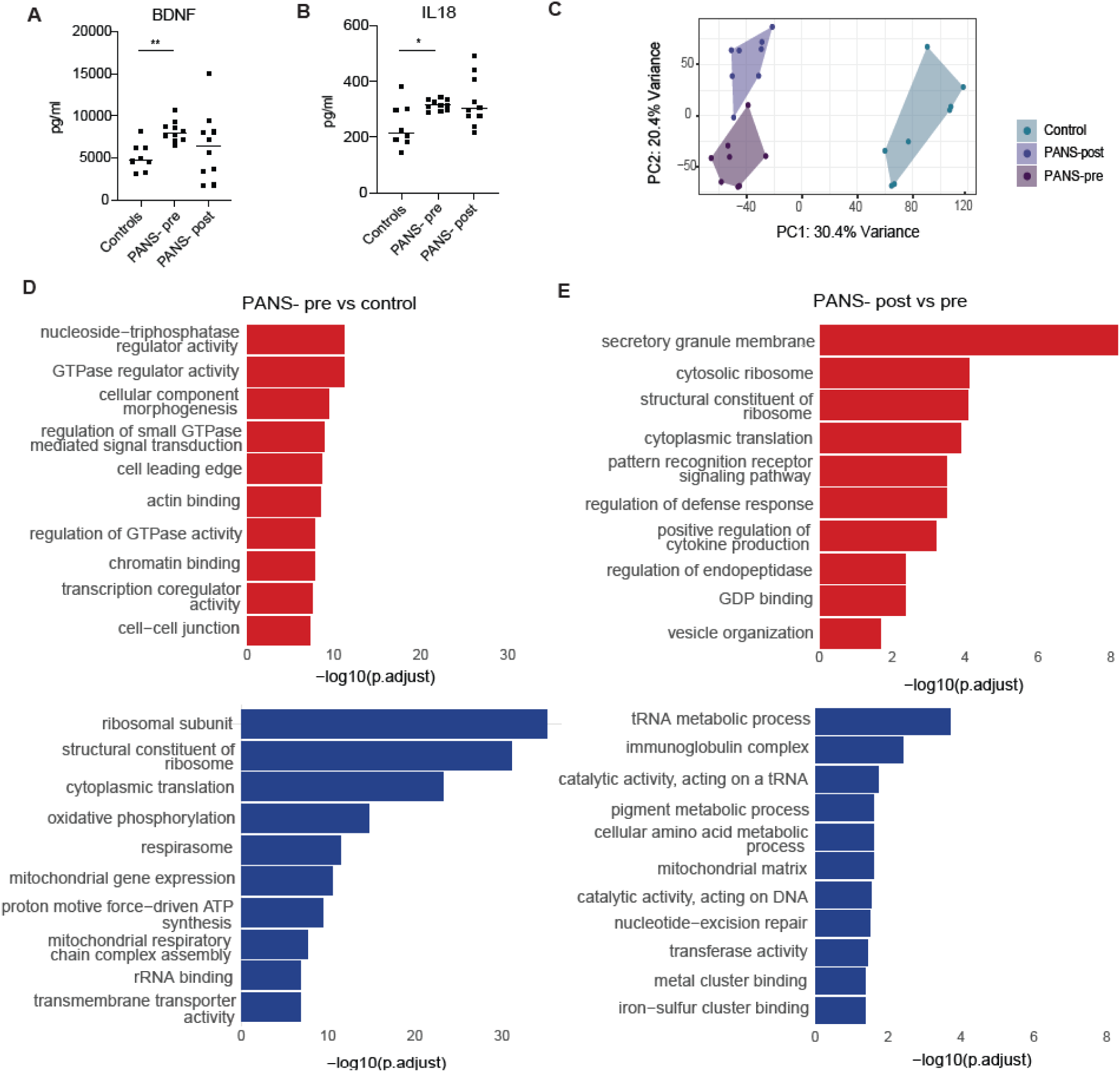
Changes in plasma immunological markers and whole blood bulk transcriptomics in children with PANS following 12 weeks of NTI164. (A) Brain-derived neurotrophic factor (BDNF) expression levels and (B) interleukin-(IL-)18 expression levels are increased in plasma of children with PANS-pre compared to healthy controls. Analysed with Wilcoxon matched-pairs signed rank test, n = 10, *P < 0.05, **P < 0.01. Data are presented as mean, but analysed with the Wilcoxon matched-pairs signed rank test due to non-normal distribution. (B) Principal component analysis (PCA) performed on bulk RNA sequencing shows clear discrimination between healthy controls, PANS-pre and PANS-post. The x-axis represents Principal Component 1 (PC1), while the y-axis represents Principal Component 2 (PC2). (C) Bar plot of GSEA GO pathways of PANS-pre vs control comparison. The top 10 up-regulated pathways (red) are related to cellular signalling (e.g. GTPase, actin), chromatin binding, and transcription related pathways. The top 10 down-regulated pathways (blue) are related to ribosome formation, cytoplasmic translation, mitochondrial function, and oxidative phosphorylation. (D) Bar plot of GSEA GO pathways of PANS-post vs pre comparison. Ribosomal and translational pathways down-regulated at baseline (PANS-pre vs control, blue) showed up-regulation after NTI164 treatment (PANS-post vs pre, red). Immune pathways, however, showed a combination of up- regulation (‘pattern recognition receptor signaling’, ‘regulation of defense response’), and down- regulation (‘immunoglobulin complex’) after NTI164 treatment (PANS-post vs pre).

#### Bulk-RNA sequencing

Bulk-RNA sequencing was performed in 8 patients at baseline (PANS-pre) and after 12 weeks of NTI164 treatment (PANS-post; mean age 11.4 (range 4-17) years), 37.5% females), and 8 age- and sex-matched controls (mean age 11.8 (range 7-17) years, 37.5% females).

##### Principal component analysis and differentially expressed genes

Post RUV normalisation (Supplementary Figure 3A), principal component analysis (PCA) of bulk RNA sequencing showed clear discrimination between healthy controls, PANS-pre, and PANS-post (Figure 2C). In the PANS-pre vs control, there were 7,710 differentially expressed genes (DEGs) FDR <0.05, with 4,028 DEGs up-regulated and 3,682 down-regulated. In PANS-post vs PANS-pre, there were 4,974 DEGS FDR <0.05, with 2,794 up-regulated and 2,180 down-regulated.

##### Pathway analysis

In PANS-pre vs control, top 10 up-regulated GSEA GO pathways included cellular signalling (GTPase-related activities, ‘actin binding’), chromatin binding, and transcription-related pathways (Figure 2D, red). Top 10 down-regulated pathways related to ribosome formation, cytoplasmic translation, mitochondrial function, and oxidative phosphorylation (Figure 2D, blue).

After NTI164 treatment, the ribosomal and translational pathways (e.g. ‘structural constituent of ribosome’, ‘cytoplasmic translation’) that were down-regulated at baseline (PANS-pre vs controls), were up-regulated (PANS-post vs pre) (Figure 2E). Additionally, several immune pathways (‘pattern recognition receptor signalling pathway’, ‘regulation of defense response’ and ‘positive regulation of cytokine production’) were up-regulated after NTI164 treatment (PANS-post vs pre). However, other immune pathways (‘immunoglobulin complex’) were down-regulated after NTI64 treatment (PANS- post vs pre), supporting broad immune modulatory effects of NTI164.

#### Single-cell RNA sequencing

Based on the findings from bulk RNA sequencing, a deeper investigation into cell-specific gene expression patterns was conducted via single-cell RNA sequencing (scRNA-seq). We performed scRNA-seq in 4 children with PANS at baseline (PANS-pre) and after 12 weeks of NTI164 treatment (PANS-post) (mean age 15.2 (range 14-17) years, 50% females), and 4 age- and sex-matched healthy controls (mean age 14.5 years (range 12-17) years, 50% females).

##### Uniform manifold approximation and projection (UMAP) and differentially expressed genes

Based on cell markers, 11 distinct cell types were identified (Figure 3A). Neutrophils constituted the largest proportion of cell type across samples. There were no significant differences observed in cell distribution between healthy controls, patients at baseline (PANS-pre), and after treatment (PANS- post) (Supplementary Figure 4B). In PANS-pre vs control, DEGs (FDR <0.05) were predominantly down-regulated, most notably in neutrophils, monocytes, and CD4+ T cells (Figure 3B, left). In PANS- post vs pre, DEGs (FDR <0.05) were predominantly up-regulated, most notably in neutrophils, monocytes, CD8+ T cells, CD4+ T cells, and B cells (Figure 3B, right).

**Figure 3.**
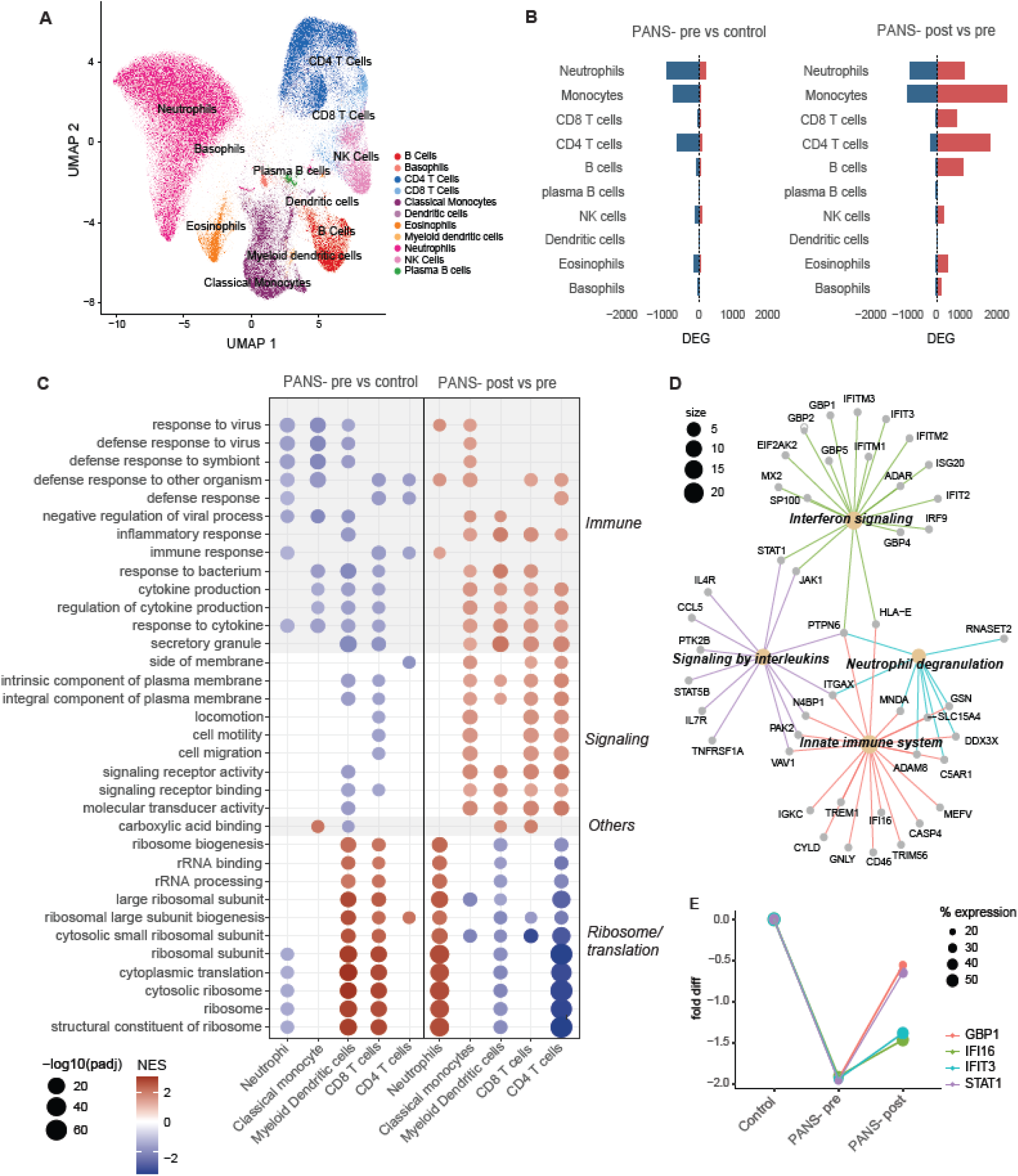
Single-cell RNA sequencing shows immune and epigenetic dysfunction in PANS patients, which was modified by NTI164. (A) Uniform manifold approximation and projection (UMAP) of all 12 samples, including 4 PANS patients before and after NTI164 treatment, and 4 controls, of single-cell transcriptomics identified 11 distinct cell types: B cells, basophils, CD4+ T cells, CD8+ T cells, classical monocytes, dendritic cells, eosinophils, myeloid dendritic cells, neutrophils, natural killer (NK) cells, and plasma B cells. (B) Bar charts of proportions of significantly differentially expressed genes (DEGs) across cell types in PANS-pre vs controls (left), and PANS-post vs pre (right). Significant DEGs were seen in neutrophils, monocytes, CD8+ T cells, CD4+ T cells, and B cells. Colour indicates direction of change (blue = down- regulated, red = up-regulated). (C) Dot plot of GSEA GO top 10 pathways in five representative cell types, neutrophils, classical monocytes, myeloid dendritic cells, CD8+ T cells, and CD4+ T cells, in PANS-pre vs control (left column), and PANS-post vs pre (right column). In PANS-pre vs control, pathways in immune function and cell signaling were broadly down-regulated across all cell types, whereas ribosomal and RNA processing pathways showed cell-specific dysregulation: down-regulation in neutrophils but up- regulation in myeloid dendritic cell and CD8+ T cells. After NTI164 treatment, the expression patterns in these pathways were reversed (PANS-post vs pre). Size of circle corresponds to degree of significance, colours indicate direction of change (blue = down-regulated, red = up-regulated). (D) Connectivity network (CNET) plot of the classical monocyte ‘defense response to other organism’ pathway. We focussed on the 69 genes that were down-regulated in PANS-pre vs control and up- regulated in PANS-post vs pre. Subcluster analysis via GO Molecular Function revealed themes in ‘interferon signalling’, ‘signaling by interleukins’, ‘neutrophil degranulation’, and ‘innate immune system’. (E) Fold change differences in the expression of key genes (*GBP1, IFI6, IFIT3, STAT1*) enriching the neutrophil ‘defense response to other organisms’ pathway in PANS-pre vs control comparison. Gene expression was down-regulated in PANS-pre compared to controls and increased after NTI164 treatment (PANS-post). The size of the dot correlates to the percentage of neutrophils expressing the corresponding gene.

##### Pathway analysis

The most significantly enriched GSEA GO pathways were identified in neutrophils, classical monocytes, myeloid dendritic cells, CD8+ T cells, and CD4+ T cells (Figure 3C). In PANS-pre vs controls, top down-regulated GSEA GO pathways were in immune function (‘defense response to virus/other symbiont/bacterium’, ‘cytokine production’, ‘inflammatory response’) and cellular signalling (Figure 3C, left column). Additionally, ribosomal and RNA processing pathways showed cell-specific dysregulation in PANS-pre vs control, with down-regulation in neutrophils but up- regulation in myeloid dendritic cells and CD8+ T cells.

After NTI164 treatment, the immune function and cellular signalling pathways that were down- regulated at baseline (PANS-pre vs control), were generally up-regulated (PANS-post vs pre) (Figure 3C, right column). The translation and ribosomal pathways that were dysregulated at baseline (PANS-pre vs controls) also demonstrated reversal after NTI164 treatment (PANS-post vs pre), most evident in neutrophils and myeloid dendritic cells.

We focused on the 69 genes from the classical monocyte ‘defense response to other organism’ pathway that were significantly down-regulated (total 119 genes) in PANS-pre vs control, and up- regulated in PANS-post vs pre (total 143 genes) (Figure 3D). Connectivity enrichment plot (CNET) of the 69 overlapping genes revealed GO molecular function subclusters, including interferon signaling (enriched by interferon-related genes: *IFIT2, IFIT3, IFITM1, IFITM2, IFITM3*), signaling by interleukins (enriched by signal transducer and activator of transcription genes: *STAT1, STAT5B,* Janus kinase genes: *JAK1)*), neutrophil degranulation and innate immune system (enriched by human leukocyte antigen genes: *HLA-E*, complement genes: *C5AR1,* integrin genes*: ITGAX*).

##### Gene expression of key genes

We further investigated trends of gene expression in key genes within the neutrophil GSEA GO pathway ‘defense response to other organisms’, including *GBP1, IFI6, IFIT3,* and *STAT3* (Figure 3E). These genes were significantly down-regulated in PANS-pre (compared to control), and increased in PANS-post, although not completely normalised to the level of healthy controls.

### PANS children have dysregulated immune and epigenetic signatures at a proteomic and phosphoproteomic level, which were modified by NTI164

#### Proteomics

Bulk proteomics was performed on peripheral blood mononuclear cells (PBMCs) from 6 children with PANS at baseline (PANS-pre) and after 12 weeks of NTI164 treatment (PANS-post) (mean age 12 (4–17) years, 33.3% females), and 6 age- and sex-matched controls (mean age 12.3 (7–17) years, 33.3% females).

##### Differentially expressed proteins

Post RUV normalisation, principal component analysis (PCA) of bulk proteomics showed clear discrimination between healthy controls, PANS-pre, and PANS-post (Supplementary Figure 5A). In PANS-pre vs control, there were 4,518 differentially expressed proteins (DEPs) FDR <0.05, with 2,349 significantly up-regulated DEPs (Figure 4A, red) and 2,169 down-regulated DEPs (Figure 4A, blue). In PANS-post vs pre there were 3,502 DEPs FDR <0.05, with 1,654 up-regulated DEPs and 1,848 down- regulated DEPs.

**Figure 4.**
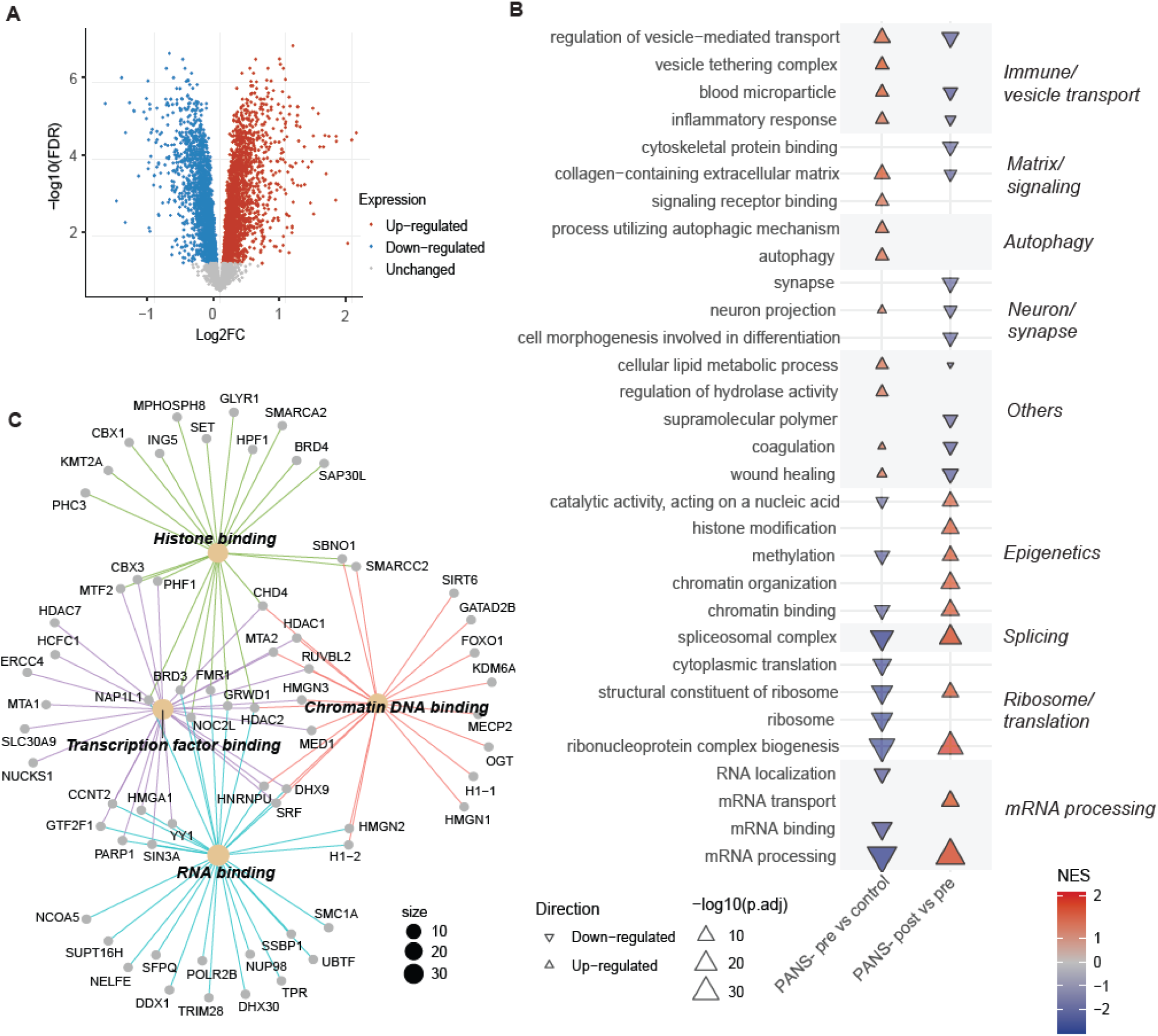
Immune and epigenetic dysregulation in the proteome of PANS children is modifiable with NTI164. (A) Volcano plot of 894 differentially expressed proteins (FDR <0.05), 341 were down-regulated (blue), and 553 up-regulated (red). (B) Arrow plot of GSEA GO top 10 most up- and down- regulated pathways in PANS-pre vs controls (left), and PANS-post vs pre (right). In PANS-pre vs control, up-regulated pathways relate to immune function, signaling, and autophagy, while down-regulated pathways relate mRNA processing, ribosome/translation, and epigenetics. These pathways were largely reversed in the PANS-post vs pre comparison. The colour of the triangle depicts the normalised enrichment score (NES), with blue indicating down-regulation, red indicating up-regulation, and no significant value indicated by white. The size of the triangle corresponds to the −log10(padj value) of the pathway. (C) Connectivity network (CNET) plot of proteins enriching the ‘chromatin binding’ pathway. We focussed on the 87 overlapping proteins that were down-regulated in PANS-pre vs control and up- regulated in PANS-post vs pre. Subcluster analysis via GO Molecular Function-related revealed themes in ‘histone binding’, ‘DNA binding’, ‘transcription factor binding’, and ‘RNA binding’.

##### Pathway analysis

In PANS-pre vs control, top 10 up-regulated GSEA GO pathways were related to immune function, signaling, and autophagy (Figure 4B, left column, red). Top 10 down-regulated pathways were related to mRNA processing, ribosome/translation, epigenetics (‘chromatin binding’, ‘methylation’) (Figure 4B, left column, blue). NTI164 treatment considerably modified protein expression in many pathways (Figure 4B, right column). Immune pathways that were up-regulated at baseline (PANS-pre vs control) showed significant down-regulation post treatment (PANS-post vs pre). Additionally, epigenetic and ribosome/translational pathways which were down-regulated at baseline (PANS-pre vs control) became significantly up-regulated after treatment (PANS-post vs pre).

We further investigated the 87 overlapping proteins from the ‘chromatin binding’ pathway that were significantly down-regulated in PANS-pre vs control (total 129 proteins), and up-regulated in the PANS-post vs pre (total 121 proteins) (Figure 4C). CNET of the 87 overlapping proteins revealed GO molecular function subclusters, including histone binding (enriched by histone deacetylases: *HDAC2,* lysine methyltransferase: *KMT2A,* epigenetic regulators: *BRD3, BRD4*), transcription factor binding (enriched by histone deacetylases: *HDAC1*, *HDAC7*), chromatin DNA binding (enriched by lysine demethylase: *KDM5A,* histone genes*: H1-1,* epigenetic regulator: *MeCP2,* transcription factor*: FOXO1*), and RNA binding (enriched by histone genes: *H1-2*).

#### Phosphoproteomics

Bulk phosphoproteomics was performed on the same samples which underwent proteomics analysis (n = 6 patients pre and post and n=6 controls).

##### Differentially abundant phosphopeptides

Post RUV normalisation, principal component analysis (PCA) of bulk phosphoproteomics showed clear discrimination between healthy controls, PANS-pre, and PANS-post (Supplementary Figure 6A). In the PANS-pre vs control, there were 4,928 differentially abundant phosphopeptides (FDR <0.05), with 2,517 significantly up-regulated and 2,411 down-regulated differentially abundant phosphopeptides. In the PANS-post vs pre comparison there were 4,271 differentially abundant phosphopeptides (FDR <0.05), with 2,103 up-regulated and 2,168 down-regulated differentially abundant phosphopeptides.

##### Pathway analysis

In PANS-pre vs control, top 10 ORA GO up-regulated pathways were cadherin binding, actin and protein kinase pathways (Figure 5A, left column, red). The top 10 ORA GO down-regulated pathways were GTPase-related activity, chromatin (‘chromatin binding’, ‘mRNA processing’), and transcription pathways (Figure 5A, left column, blue).

**Figure 5.**
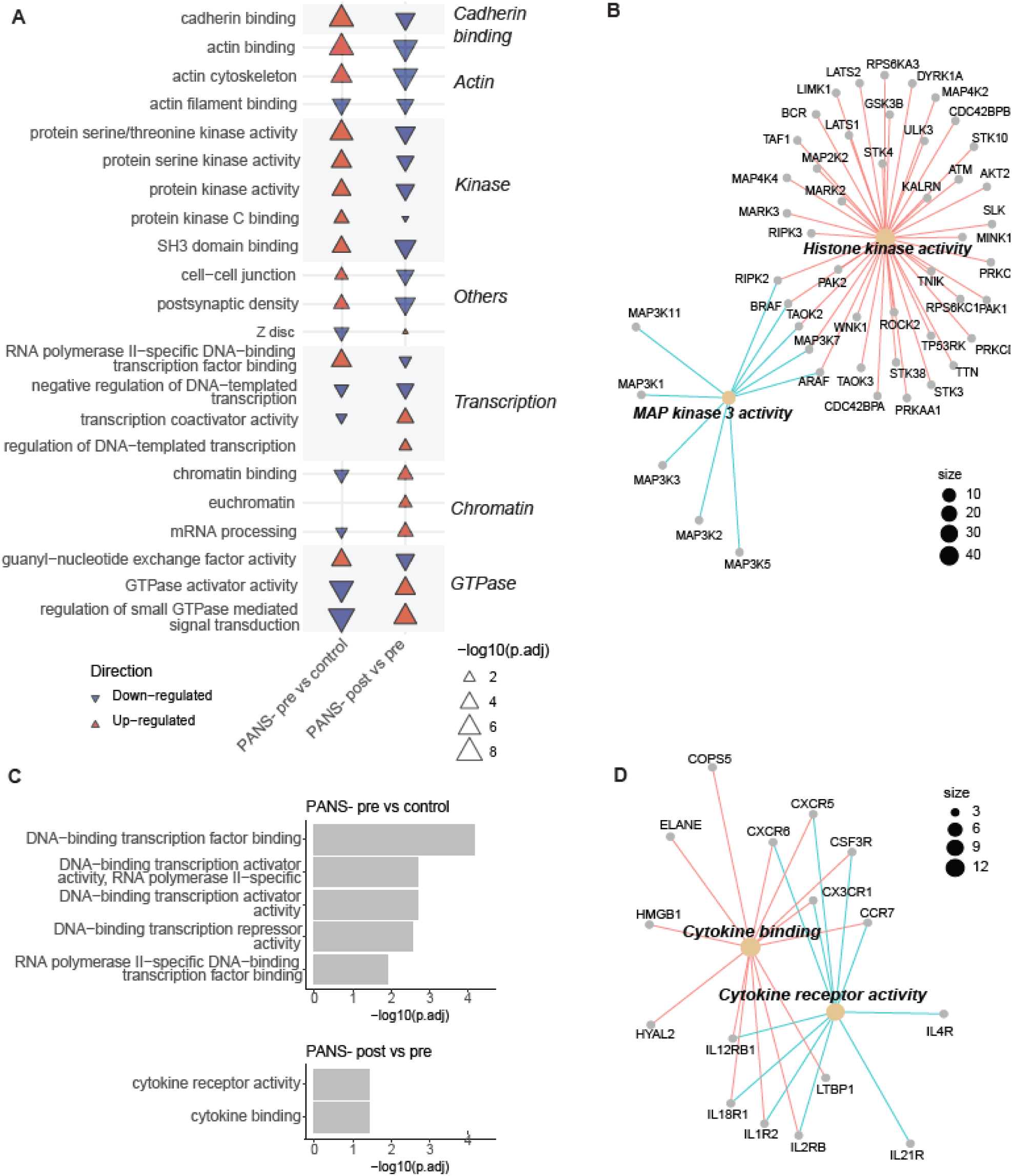
Immune and epigenetic dysregulation in the phosphoproteome and DNA methylome of PANS children is modifiable with NTI164. (A) Arrow plot of ORA GO top 10 most up- and down-regulated pathways in PANS-pre vs controls (left), and PANS-post vs pre (right). In PANS-pre vs control, up-regulated pathways relate to cadherin binding, actin and kinase activity, and down-regulated pathways relate to GTPase signaling, chromatin and transcription. The baseline dysregulation in these pathways is significantly modified by NTI164 (PANS-post vs pre). Blue indicates down-regulation, and red indicates up-regulation, and no significant value indicated by white. The size of the triangle corresponds to the −log10(padj value) of the pathway. (B) Connectivity network (CNET) plot of ORA GO ‘protein serine/threonine kinase activity’. We focussed on the 53 proteins enriched by the overlapping phosphopeptides that were up-regulated in PANS-pre vs control and down-regulated in PANS-post vs pre. A subcluster analysis via GO Molecular Function revealed themes in ‘histone kinase activity’ and ‘MAP kinase 3 activity’. (C) Bar chart of ORA molecular function top 5 differentially methylated region pathways in the PANS- pre vs control comparison (top), and the 2 differentially methylated region pathways in the PANS- post vs PANS-pre comparison (bottom). All pathways in PANS-pre vs control comparison related to DNA transcription, and pathways in PANS-post vs pre related to cytokine activity. (D) CNET of the cytokine activity pathways that were dysregulated in PANS-post vs pre DNA methylation analysis revealed chemokine and interleukin genes corresponding to differentially methylated regions (Figure 5D).

Modification of these pathways was seen following 12 weeks of NTI164 administration (PANS-post vs pre, Figure 5A, right column). The cadherin binding, actin, and protein kinase activity pathways that were up-regulated in patients at baseline (PANS-pre vs control) were down-regulated after NTI164 treatment (PANS-post vs pre). Additionally, many GTPase signalling, chromatin, and transcription pathways that were down-regulated at baseline (PANS-pre vs control) were up- regulated following NTI164 treatment (PANS-post vs pre).

We focused on the 53 proteins enriched by phosphopeptides from the ‘protein serine/threonine kinase activity’ pathway that were significantly up-regulated in the PANS-pre vs control comparison (74 proteins), and down-regulated in the PANS-post vs pre comparison (62 proteins) (Figure 5B).

CNET of these 53 overlapping proteins enriched by phosphopeptides revealed GO molecular function subclusters, including histone kinase activity, and MAP kinase 3 activity.

### PANS children have differential methylation in DNA transcription factor regions, and NTI164 has DNA methylation effects on immune pathways

DNA methylation was performed on whole blood from 8 children with PANS at baseline (PANS-pre) and after 12 weeks of NTI164 treatment (PANS-post; mean age 11.4 (range 4-17) years, 37.5% females), and 8 age- and sex-matched controls (mean age 11.8 (range 7-17) years, 37.5% females).

#### Differentially methylated positions

Post RUV normalisation, principal component analysis (PCA) of DNA methylation showed discrimination between healthy controls, PANS-pre, and PANS-post (Supplementary Figure 7D). In the PANS-pre vs control, there were 63,749 differentially methylated positions (FDR <0.05). In the PANS-post vs pre comparison there were 17,930 differentially methylated positions (FDR <0.05).

#### Differentially methylated region pathway analysis

Differentially methylated positions were assigned to differentially methylated regions. In PANS-pre vs control, top 5 significant ORA GO MF dysregulated pathways were all related to DNA transcription and included *HDAC* genes and *TBX* genes (Figure 5C, top). Following 12 weeks of NTI164 administration (PANS-post vs pre, Figure 5C, bottom), the 2 significant ORA GO Molecular Function dysregulated pathways involved cytokine activity. A CNET of these cytokine activity pathways revealed chemokine and interleukin genes corresponding to differentially methylated regions (Figure 5D).

## DISCUSSION

This is the first open-label clinical trial of full-spectrum medicinal cannabis in children with PANS, integrating clinical outcomes with multi-omics analyses to evaluate the efficacy of NTI164. Based on our hypothesis that PANS is an epigenetic disorder of immune and brain function, we selected a cannabis-based treatment due to its recognised epigenetic and immune modulating effects. NTI164 was well-tolerated at doses up to 20mg/kg/day, with only one serious adverse event (diarrhoea) reported during the trial, and the child and family chose to remain on the drug. We observed significant clinical and functional improvements in children with PANS following treatment with NTI164. Using a multi-omics approach, we also identified dysregulated epigenetic (chromatin, methylation, histone modifications, transcription factor), ribosomal, mRNA processing/transcriptional, immune, and signaling (GTPase, actin) pathways in children with PANS, and showed that this dysregulation was modifiable with NTI164. We previously identified an immune, ribosomal, and epigenetic signature in peripheral immune cells from children with PANS and neurodevelopmental regression in children who lacked highly penetrant DNA variants (10). This abnormal signature was reversed following treatment with the immunomodulator intravenous immunoglobulin (IVIg) (17, 18). Here, we validated our previous baseline findings in children with PANS and demonstrated similar therapeutic benefits using a proprietary strain of medicinal cannabis (NTI164).

Prior to treatment with NTI164, the PANS patients in our cohort were very debilitated with substantial functional impairment. All patients had previously trialled multiple medications and conventional therapy, and remained seriously functionally impaired. Additionally, the children suffered ongoing infection-triggered relapses which exacerbated impairments. Improvement in emotional assessments was the primary outcome of this trial. Following treatment with NTI164, clinical assessments demonstrated improvement in emotional regulation, anxiety, OCD, ADHD, tics, and overall Quality of Life (Figure 1, Supplementary Figure 1, Supplementary Table 4). There were significant improvements in social phobia, panic disorder, major depression, separation anxiety, generalised anxiety, and obsessive-compulsive symptoms, as assessed by the RCADS-P. A previous study of NTI164 in ASD also showed similar clinical improvements in areas of anxiety, emotional regulation, and attention (19).

We incorporated a multi-omics approach to explore the *ex vivo* biological effects of NTI164. In this study of children with PANS, we observed abnormal gene and protein expression at baseline, along with altered DNA methylation, enriched in pathways related to immune system function and immune activation, which were modulated by NTI164 treatment. Based on single-cell RNA sequencing, we observed down-regulation of immune pathways in multiple cell types in PANS, affecting both adaptive and innate immune cell types, which were reversed with NTI164 treatment. Immune system dysfunction in PANS children pre-NTI164 was also supported through cytokine testing, which showed trends of elevated expression levels in cytokines in PANS pre-NTI164 compared to controls, and trends of reduced cytokine values post-NTI164. IL-18 and BDNF were also found to be significantly elevated in PANS children at baseline compared to healthy controls. The role of BDNF in CNS development and synaptic plasticity is well known (20, 21), and a previous study in ASD patients suggests a link between increased BDNF levels and intellectual disability (22). Although the cytokine and BDNF findings added some insights, these approaches were statistically less powerful compared to the RNA/proteomic approaches.

Cannabis, and in particular the isolated cannabinoids CBD and THC, has demonstrated clinical benefit in various immune and autoimmune related neurological conditions, including multiple sclerosis, neuropathic pain, and neurodegenerative conditions, in part due to its potent anti-inflammatory effects and reduction of oxidative stress (23). Cannabinoid receptor CB2 is predominantly expressed on immune cells and interaction between the receptor and various cannabinoids can suppress inflammatory cytokine production, and alter T cell differentiation, macrophage activity, and B cell function (24–26). NTI164 has similarly been shown to suppress production of inflammatory cytokines and reduce expression of microglia activation state markers (15).

Although we focus on peripheral immune cells in this study, microglia, which are resident immune cells within the central nervous system, are hypothesised to be chronically activated in PANS (27). Disrupted microglia-dependent synaptic pruning is a key process in the pathophysiology of NDDs (28), and epigenetic ‘priming’ of microglia and peripheral immune cells can lead to widespread immune system dysregulation (29, 30). A ‘two-hit’ model has been proposed for the development of NDDs, in which prenatal disruptions, such as maternal immune activation and chronic inflammation *in utero*, prime the foetal brain (8, 31). A subsequent postnatal ‘second hit’ postnatally such as infection or psychosocial stress, is thought to trigger immune dysregulation, leading to aberrant microglial activation and the clinical manifestation of NDDs (27). We found high rates of maternal autoimmunity in our cohort, which has been reported in other PANS cohorts (31), and represents an example of the genetic and environmental (immune activation in pregnancy) factors relevant to PANS expression (8). Maternal autoimmunity is known to be associated with increased expression of neurodevelopmental disorders (ASD, ADHD, Tourette) in offspring (8, 32). CBD is known to modulate an extensive array of microglial genes involved in regulating stress and inflammation (33), ameliorating microglia-dependent inflammation (34–36), and limiting neuronal damage in excitotoxic environments (37). *In vitro* studies of NTI164 have shown attenuation of microglial activation and associated pro-inflammatory cytokines, and reduced neuronal cell death following a stressor in cells treated with NTI164 (15), and these may be mechanisms through which our observed clinical dysregulation is normalised.

Secondly, we observed ribosomal and translational dysregulation in children with PANS compared to healthy controls through RNA and proteomic sequencing. Interestingly, the direction of dysregulation of translation-related pathways varied across cell types. For instance, ‘structural constituent of ribosome’ was down-regulated at baseline in neutrophils, and up-regulated in myeloid dendritic cells and CD8+ T cells. There are well-documented links between translation and ribosomal protein abnormalities in NDDs (38–40). Ribosomal function is fundamental for maintaining cellular homeostasis, through translation and regulation of protein synthesis. Disruption to ribosome biogenesis can reduce synthesis of proteins critical for dendritic and synaptic function, contributing to changes in neuronal function and connectivity in NDDs (41). Additionally, ribosomal proteins can be involved in immune function (42). Ribosome stress can trigger production of ribosomal proteins involved in signalling pathways, potentially contributing to downstream immune system dysregulation observed in PANS (17).

Thirdly, we identified broad epigenetic dysregulation relating to chromatin, methylation, histone modification and transcription factors at baseline across all omics methodologies, which was significantly modified with NTI164 treatment. We also identified dysregulation of GTPase and actin cytoskeletal pathways in children with PANS, which were reversed with NTI164 treatment. Increasing evidence supports the role of GTPase signaling and actin dynamics in chromatin remodelling, suggesting that they are key mechanisms within epigenetic control (43, 44). Epigenetic dysregulation plays a crucial role in onset and progression of various NDDs (17, 45). Epigenetic mechanisms such as chromatin modification, DNA methylation, and transcription factors are essential for regulating gene expression and disruptions in these processes can lead to abnormal neural differentiation and connectivity. Chromatin abnormalities have been implicated in NDDs, and chromatin structure tightly regulated by histone modifications (46, 47). Proteomics analysis in our current study revealed proteins contributing to chromatin binding, specifically histone deacetylases, lysine demethylases, lysine methyltransferases, which were down-regulated at baseline and up- regulated with NTI164. Our study showed that protein kinases with histone kinase activity were abnormal in PANS children and modified by NTI164; histone phosphorylation by kinases is also involved in chromatin structure and function (48). Further research is needed to explore the effects of NTI164 on histone kinases and phosphorylation of histones, as well as downstream effects on chromatin structure and gene expression, plus other histone modification (methylation, acetylation etc).

Analysis of DNA methylation, another epigenetic modification, in our current study revealed PANS patients have differences in methylation in DNA transcription-related regions, supporting the idea that PANS is an epigenetic disorder with associated abnormal gene expression and downstream effects on the immune system. Some studies suggest methylation changes in genes relating to synaptic or neuronal development leads to altered brain function contributing to OCD, anxiety, and tics (49, 50). Our study showed that NTI164 has effects on DNA methylation pathways relating to immune function.

The epigenetic effects of cannabis are only beginning to be explored (51, 52). Isolated CBD can affect histone acetyltransferases and histone deacetylases, and a study in adult mice demonstrated CBD can also work synergistically with other cannabinoids to induce histone modifications in the mesolimbic system and improve motor outcomes and anxiety (53). Cannabis’ effect on epigenetic ‘reprogramming’ has also been suggested to involve repression of genes relating to inflammation and thus dampening immune responses, and promoting immunosuppressive effects (54). Cannabinoids bound to CB1 and CB2 receptors can inhibit transcription factors associated with modification of inflammatory gene expression (25, 55), and certain histone modifications induced by THC and CBD have been shown to promote heterochromatin formation and gene ‘silencing’, leading to long-term suppression of specific transcripts (56, 57). More research into the epigenetic effects of cannabis and NTI164 is required, utilising standardised drug compositions and assessments. This should include analysis of the effects of NTI164 on histone modifications, chromatin function, and methylation, and its downstream effects on expression of NDD and immune genes. Chromatin analyses (e.g. ATAC-seq or ChIP-seq) and microRNA analysis would be useful in further exploring epigenetic dysregulation in PANS (58).

Limitations of the current study include the small sample size and open-label nature of our clinical trial. As this was a Phase I/II, proof-of-concept trial, an open-label design was pursued. A Phase II/III, randomised, placebo-controlled, double-blind clinical trial of NTI164 in PANS is currently under development. Our patients were also on various other medications, including anti-psychotics and anti-depressants, which could influence baseline RNA or protein expression, but we controlled for this by prohibiting changes to medication regimes in the three months prior to enrolment and during the study period. Also, our multi-omics investigations were based on peripheral immune cells and not central nervous system cells, however emerging preclinical research does show comparable dysregulation between the periphery and brain tissue in an animal model of NDDs (59). Further investigation using animal models or patient-derived induced pluripotent stem cells (iPSCs) would be useful to determine the effects of NTI164 on brain cells in the context of PANS. Additional studies are also needed to further validate our findings, including functional immune testing, ribosomal profiling, epigenetic and phosphorylation studies, and to investigate the long-term effects and safety of cannabis.

## Conclusion

This study demonstrates the clinical benefits of NTI164, a novel full-spectrum medicinal cannabis plant extract, in children with PANS. Our results show NTI164 has epigenetic, ribosomal and immune modulatory effects, and can reverse baseline abnormalities observed in PANS. While further studies are needed, these results show significant potential for NTI164 in PANS and potentially other neurological and epigenetic conditions involving immune dysregulation.

## METHODS

### Ethics approval

Ethics approval for this study was granted by the Sydney Children’s Hospitals Network (SCHN) Human Research Ethics Committee (HREC reference: 2022/ETH02308) and Monash Health (HREC reference: RES-23-0000-333X).

### Participant selection

Children were recruited from specialist neurology clinics at the Children’s Hospital at Westmead (CHW, clinic run by RD) or Monash Children’s Hospital (MCH, clinic run by MF and MK). All children were diagnosed with PANS according to the criteria from Swedo et al. (1), and Chang et al. (16) and also met eligibility criteria for the current study (for full inclusion/exclusion criteria, see Supplementary material, Appendix 1).

All participants and their parent/guardian provided written informed consent for participation in this study.

Age- and sex- matched healthy control children (children of hospital staff members) were recruited for biological investigations. The inclusion criteria for controls were the absence of neurodevelopmental disorders, autoimmune conditions, moderate-severe allergies, and no recent infections (within 2 weeks of blood sample collection).

### Structured clinical assessment on REDCap and clinician-directed interview (questionnaires)

A structured clinical interview was conducted between parents of participants and trial staff to gather information regarding family background and familial medical history and symptoms at the time of recruitment. This allowed family medical history and information relating to patient symptoms to be collected in a standardised manner across participants and sites.

### Study design

This study was split into 4 stages: up-titration, treatment, down-titration, and extension. In the up-titration phase, patients received a starting dose of NTI164 of 5mg/kg/day. This was increased by 5mg/kg weekly for 4 weeks, until the target dose of 20mg/kg/day (or maximum tolerated dose as determined by the treating physician) was reached by Week 4 of the study.

In the treatment phase, children received NTI164 at 20mg/kg/day (or their maximum tolerated dose) for 8 weeks. Blood samples were collected at Week 12 (i.e. end of the treatment phase, primary endpoint), as well as questionnaires completed.

At Week 12, children wishing to stop NTI164 were down-titrated by 5mg/kg/week from 20mg/kg/day (i.e. reversal of up-titration), and Week 16 was the end of the study for these children.

At Week 12, children wishing to remain on NTI164 and enter the extension phase were able to do so.

### Questionnaires

Questionnaires used in this study are gold-standard tools, see Supplementary Table 3 for more detail on individual questionnaires. Questionnaires were administered by trained personnel at baseline (i.e. prior to NTI164), at Week 4 (i.e. completion of up-titration phase/start of treatment phase), and at Week 12 (i.e. completion of treatment phase). Parent/caregiver-rated questionnaires were completed at the same time points in the presence of study staff.

### Sample collection

#### Clinical Pathology

After written consent, venous blood was collected prior to starting NTI164 (baseline) as well as after 12 weeks of receiving the drug. See Supplementary Table 1 for detailed information about parameters investigated.

#### Plasma neuroinflammation panel

Venous blood was collected into lithium heparin tubes and centrifuged at 1,300xG for 10 minutes at room temperature. Separated plasma was divided into 200µL aliquots and stored at −80°C until analysis.

#### Samples for bulk-RNA Sequencing

Venous blood was collected directly into PAXGene™ Blood RNA tubes and stored at −80°C until extraction and sequencing by the Australian Genome Research Facility Ltd (AGRF Ltd).

#### Samples for single-cell RNA sequencing (HIVE™ sample capture)

Single-cell RNA sequencing was performed using the HIVE™ Single Cell Solution kit from Honeycomb Biotechnologies, Inc. Venous blood was collected into acid-citrate-dextrose (ACD) tubes, and red blood cells (RBCs) were removed following the EasySep™ RBC Depletion protocol from STEMCELL Technologies (catalogue #18170) using the EasySep™ Magnet (catalogue #18000, Stemcell Technologies).

The resulting cell suspension was then loaded into the HIVE™ collectors following the manufacturer’s protocol (Honeycomb Biotechnologies, Inc., USA) and stored at −80°C until library preparation and sequencing by AGRF Ltd. Samples were always loaded into a HIVE™ collector within 1 hour of collection to reduce neutrophil activation.

#### Samples for methylation

Venous blood was collected directly into EDTA K2 tubes and stored at −80°C until analysis by AGRF Ltd.

#### Samples for proteomics and phosphoproteomics

A minimum volume of 4mL venous blood was collected into ACD tubes and diluted 1:1 with Dulbecco’s phosphate buffer saline (DPBS) −Ca2+/−Mg2+. Samples were then layered over Ficoll in SepMate™-15 tubes (STEMCELL Technologies, catalog #85415) and centrifuged at 1,200xG for 15 minutes at room temperature (RT). Supernatant was discarded and the peripheral blood mononuclear cell (PBMC) layer was transferred via pipette into a 50mL tube. Tubes were filled to 50mL with DPBS and centrifuged at 1,000xG for 6 minutes at RT, twice.

Following the second spin, cells were resuspended in 200µL lysis buffer with detergent (50mM Tris, 0.8% v/v Triton X-100, Complete EDTA-free Protease Inhibitor Cocktail (Merck), PhosSTOP (Merck)) + 2uL of 200mM phenylmethylsulfonyl fluoride (PMSF) in ethanol, transferred to a 1.5mL tube, heated at 85°C for 10 minutes, and then stored at −80°C until analysis.

### Plasma neuroinflammation panel

A multiplex assay was performed on thawed plasma from 8 children with PANS (mean age 11.4 years, 37.5% females) and 8 age- and sex- matched controls (mean age 11.8 years, 37.5% females). The LEGENDplex™ Human Neuroinflammation Panel 1 (13-plex) assay was performed according to the manufacturer’s protocol by the Westmead Cytometry Core Facility at the Westmead Research Hub (Westmead, Australia).

### Bulk blood RNA sequencing (library preparation and sequencing)

Bulk RNA sequencing was performed on whole blood from 8 children with PANS (mean age 11.4 years, 37.5% females) and 8 age- and sex- matched controls (mean age 11.8 years, 37.5% females). This workflow included RNA extraction, depletion of ribosomal RNA via hybrid capture (Illumina Ribo-Zero), and Illumina TruSeq Stranded Total RNA Library Preparation (input 200-1,000ng of Total RNA). The stranded RNA samples were sequenced on the Illumina NovaSeq X sequencing platform (2 x 150 base pairs) for a depth of 50 million paired end reads. The cleaned sequence reads were aligned against the *Homo sapiens* genome (Build version hg38), and the STAR aligner (v2.5.3a) was used to map unique reads to the genomic sequences (60).

### Single-cell RNA sequencing (HIVE, library preparation, and sequencing)

Single-cell RNA sequencing was performed on samples from 4 patients at baseline and after 12 weeks of NTI164 treatment (mean age 15.2 years, 50% females), as well as 4 age- and sex-matched controls (mean age 14.5 years, 50% females). Following a standard protocol (Honeycomb Biotechnologies, Inc., USA), cell-loaded HIVE™ devices were sealed with a semi-permeable membrane, allowing for a strong lysis solution and a hybridisation solution. After collection, beads with captured transcripts were extracted from the HIVE™ by centrifugation and transferred to a 96- well plate (AGRF Ltd., Westmead). The size profiles of the final libraries were determined on a TapeStation platform using a D5000 ScreenTape System (Agilent Technologies, Santa Clara, CA, USA), and the concentration of final pooled libraries was determined by qPCR. HIVE™ scRNAseq libraries were sequenced using specific primers contained in the kit on an Illumina® NovaSeq X sequencer (AGRF Ltd., Melbourne).

### Proteomics

Proteomics analysis was performed in 6 patients (PANS-pre and PANS-post, mean age 12 years, 33% female), as well as 6 age- and sex-matched control children (mean age 12.3 years, 33% female). The samples were lysed, digested, and tagged using the TMTpro 18-plex system (n = 6 for each of three groups). Hydrophilic ion liquid chromatography fractionation was performed on a Vanquish Neo HPLC system with a 250 mm long and 1 mm inside diameter TSKgel Amide-80 column (Tosoh Biosciences, Inc., OH, USA). Fractions were collected into a 96-well plate using an FC204 fraction collector (Gilson) at 1 min intervals, monitored by absorbance of UV at 214 nm. Fractions were analysed by LC-MS/MS, performed using a Dionex UltiMate 3000 RSLC nano system and Q Exactive Plus hybrid quadrupole-orbitrap mass spectrometer (Thermo Fisher Scientific). An in-house 300 mm long 0.075 mm inside diameter column packed with ReproSil Pur C18 AQ 1.9 μm resin (Dr Maisch, Germany) was used. The instrument settings were as described previously (61).

### Phosphoproteomics

Phosphoproteomics analysis was performed on the same samples as underwent proteomics analysis, and so all PBMC preparation was identical. Phosphopeptides were enriched and fractionated prior to LC-MS/MS analysis as described previously (62).

### Searching of LC-MS/MS data

The raw LC-MS/MS data was processed with MaxQuant v1.6.7.0 using the following settings: variable modifications were oxidation (M), acetyl (protein N-terminus), deamidation (NQ) and phospho (STY); carbamidomethyl (C) was a fixed modification; digestion was set to trypsin/P with a maximum of 3 missed cleavages; the TMTpro correction factors were entered for lots XC344112 and XK350589; minimum reporter peptide ion faction was 0.6; the *Homo sapiens* reference proteome with canonical and isoform sequences downloaded March 4 2024 with 82,485 entries and 20,597 genes; the inbuilt contaminants fasta file was also used; minimum peptide length was 6 and maximum peptide mass was 6,000 Da; second peptides search and dependent peptides searches were enabled; peptide spectrum matching and protein false discovery rates were set at 1%. All other settings were default.

### DNA methylation

Methylation analysis was performed on whole blood from 8 children with PANS (mean age 11.4 years, 37.5% females) and 8 age- and sex- matched controls (mean age 11.8 years, 37.5% females). Genome-wide DNA methylation was assessed using the Illumina MethylationEPIC v2.0 BeadChip (Illumina, Inc., San Diego, CA, USA), which interrogates >936,000 CpG sites. The BeadChips were scanned using an Illumina iScan. All analysis was undertaken using the R statistical environment (63). Quality control and probe summaries were assessed using the lumi Bioconductor package (64). All samples were retained following quality control. Differential methylation was assessed using the limma package (65).

### Bioinformatic analysis

#### Omics bioinformatic analysis

Omics data were analysed in the R statistical environment (63) with *tidyverse* (66). For bulk RNA sequencing and proteomics, filtering and normalisation steps were first performed. Subsequently, normalisation with removal of unwanted variation, via the remove unwanted variation *(RUV)* R package was performed (67). In the bulk RNA sequencing (k=13), proteomics (k=4), and phosphoproteomics (k=4) (factors of unwanted variation) were used respectively to remove genes that had minimal differential expression, compared to negative control genes. For linear modelling, the limma R package was used and the p-values were calculated using the empirical Bayes method ‘eBayes’ function (68). The false discovery rate (FDR) correction was applied to the p-values by calculating the adjusted p-values. Significant differentially expressed genes/proteins were defined as those with adjusted p-values/false discovery rate (FDR) < 0.05.

For single cell transcriptomics, the data was analysed using the Seurat package (69). Cells with a high mitochondrial transcript ratio (>0.15) were excluded. Normalisation was performed using *SCTransform* in Seurat and immune cell types were assigned with *scType* and *scPred* (70). Merged data were then split by cell type and separately normalized, scaled, integrated between patient using harmony (71), then UMAP (uniform manifold approximation and projection) projections were made using the first 30 dimensions. Differentially expressed genes with significant FDR values <0.05 were identified using *FindMarkers*.

DNA methylation profiling was performed using the Illumina Infinium MethylationEPIC v2.0 BeadChip (Illumina, Inc., San Diego, CA, USA). Raw IDAT files were imported and preprocessed using the *minfi* package (72) in R, following the Bioconductor methylation array analysis workflow (73). Low-quality samples and probes were excluded based on detection *p-values* (>0.01) and known single nucleotide polymorphisms (SNPs). Probes on sex chromosomes and non-CpG probes were also removed. Data were normalised using *preprocessQuantile*.

Methylation levels were summarised as beta and M-values, with M-values used for statistical testing. To account for unwanted technical variation, RUV-III normalisation was applied to the M-values using a control set of probes and setting unwanted factors K = 6 (74). Differentially methylated positions (DMPs) were identified using the *limma* package (68), applying empirical Bayes moderation and adjusting for multiple testing with the Benjamini-Hochberg method (FDR < 0.05).

Differentially methylated regions (DMRs) were detected using the *DMRcate* package (75), which applies kernel smoothing to identify spatially correlated methylation changes across genomic regions.

#### Pathway enrichment analysis

##### Gene Set Enrichment Analysis (GSEA) for bulk RNA seq, scRNAseq, and proteomics

The genes and proteins were ranked based on their sign(logFC) x log10Pvalue scores (76, 77). Enriched gene sets were identified based on a running sum statistic (adjusted p values) and statistical significance based on the false discovery rate (FDR). Significant GSEA Gene Ontology (GO) pathways (FDR <0.05) were further simplified use the *simplify* function in clusterProfiler, and ranked by p.adjust (78).

##### Over representation enrichment (ORA) analysis for phosphoproteomics and methylation

We took an approach that deliberately reduced the significant pathways identified by limiting the background to only proteins we could detect by mass spectrometry, and using the ranked list method within gProfiler, which weights enrichment toward highly ranked genes. We separated up- and down-regulated phosphorylation, assigned the maximum positive and negative quantitative values for phospho-regulation to each protein, and ranked the proteins using both the quantitative value and significance of maximal change.

Bar and dot plots of GSEA results were plotted using *ggplot2* package, and heatmaps of GSEA results were made using the *pheatmap* package. To further evaluate themes within pathways, gene subclusters were created based on most significant Reactome or molecular function pathways per gene set, and connectivity network (CNET) plots were created using *enrichplot*.

For methylation, probe annotations were retrieved from the *IlluminaHumanMethylation450kanno.ilmn12.hg19* annotation package. Gene ontology enrichment for DMP or DMR-associated genes was conducted using the *clusterProfiler* package, employing *enrichGO* and *enrichKEGG* functions for Gene Ontology and KEGG pathway enrichment, respectively (79). Reactome pathway analysis was also conducted using *enrichPathway* from the *ReactomePA* package (80). The same approach was used to evaluate themes within pathways using *enrichplot*.

## Data Availability

Additional data and code available upon request by qualified Investigators. Requests for further information should be directed to the corresponding author, Prof Russell Dale.

## Acknowledgements

We would like to thank the participants and their families for their contribution to this work. We would like to acknowledge the Clinical Trial Pharmacy staff at The Children’s Hospital at Westmead and Monash Health, as well as Ms Vino Dinesh and Ms Kanan Sharma from Monash Children’s Clinical Trial Centre, for their support of this trial. We also thank the Australian Genome Research Facility Ltd for their contribution to this work.

## Funding statement

This study was monetarily supported by Fenix Innovation Group Pty Ltd and Neurotech International Ltd (Sponsors). The Sponsors had no role in data collection, analysis, or presentation.

## Author contributions

BAK, MK, MCF, SP, and RCD designed the trial. BAK, VXH, HN, SSM, MK, MCF, SP, and RCD recruited the patients and controls. BAK, MK, MCF, and RCD performed clinical assessments. BAK, VXH, HN, XL, RD, SD, MG, SP, and RCD conducted experiments and acquired and analysed the data. BAK, SSM, MK, MCF, SP, and RCD oversaw the trial and data analysis. VXH, HN, NA, LLM, BSG, and MG performed computational analyses. BAK and VXH drafted this manuscript and shared responsibilities as co-first authors. SP and RCD critically revised this manuscript and shared responsibilities as co-senior authors.

## Conflict of Interest

BAK became affiliated with Fenix Innovation Group Pty Ltd (Sponsor) following completion of this trial. BAK was not affiliated with Fenix Innovation Group Pty Ltd during collection or analysis of the data presented in this paper.

All other authors declare no conflicting interests.

## Notes

### Clinical Trial

NCT06621888

### Author Declarations

Ethics approval for this study was granted by the Sydney Childrens Hospitals Network (SCHN) Human Research Ethics Committee (HREC reference: 2022/ETH02308) and Monash Health (HREC reference: RES-23-0000-333X).

