## Supplementary Figures for "Medicinal Cannabis Plant Extract (NTI164) modifies epigenetic, ribosomal, and immune pathways in paediatric acute-onset neuropsychiatric syndrome"

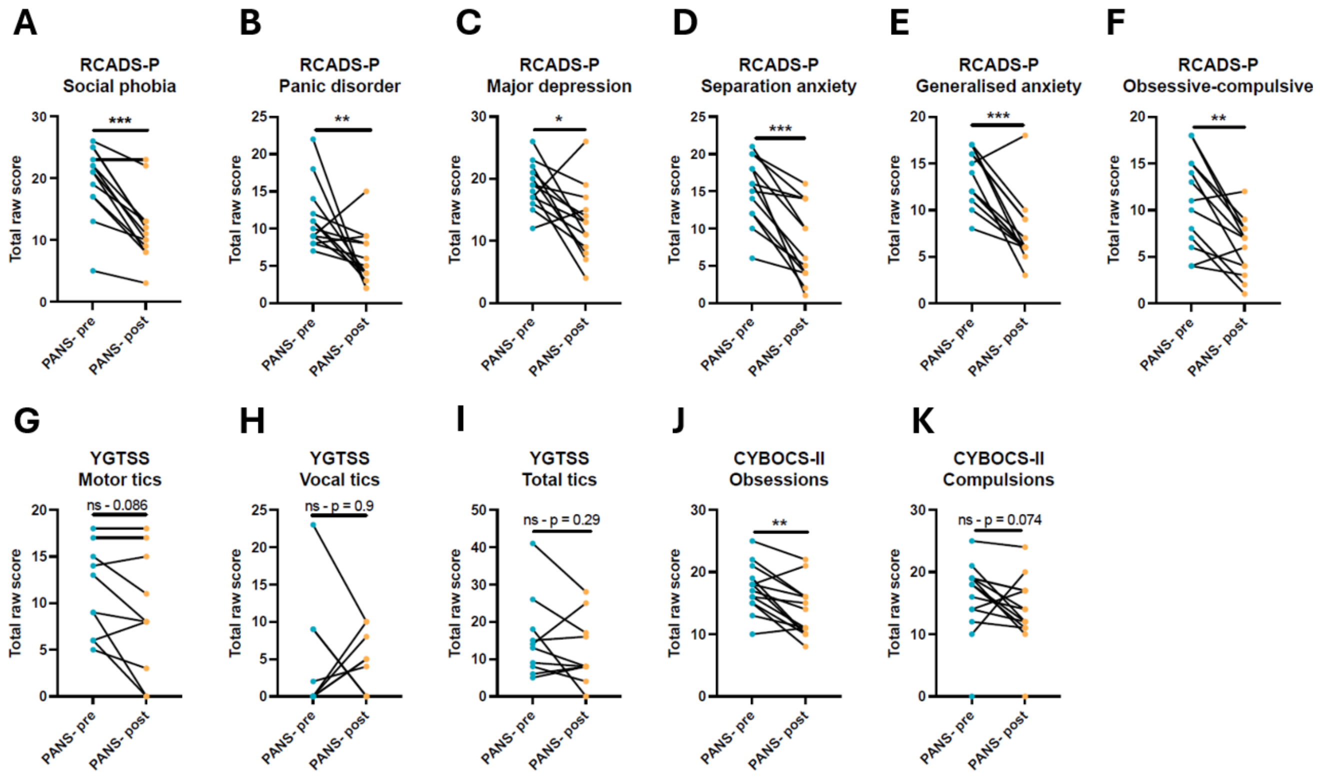


**Supplementary Figure 1. Clinical survey subdomain scores of PANS children at baseline and following 12 weeks of NTI164 administration.** Specific subdomains for gold-standard behavioural assessment scales were significantly improved following 12 weeks of NTI164 administration.

(A-F) RCADS-P subdomains: social phobia, panic disorder, major depression, separation anxiety, generalised anxiety, and obsessive-compulsive symptoms, respectively.

(G-I) YGTSS subdomains: motor tics, vocal tics, total tics, respectively.

(J-K) CYBOCS-II subdomains: obsessions, compulsions, respectively.

Wilcoxon matched-pairs signed rank test, n = 14, *P < 0.05, **P < 0.01, ****P < 0.0001.

**Supplementary Figure 2.**

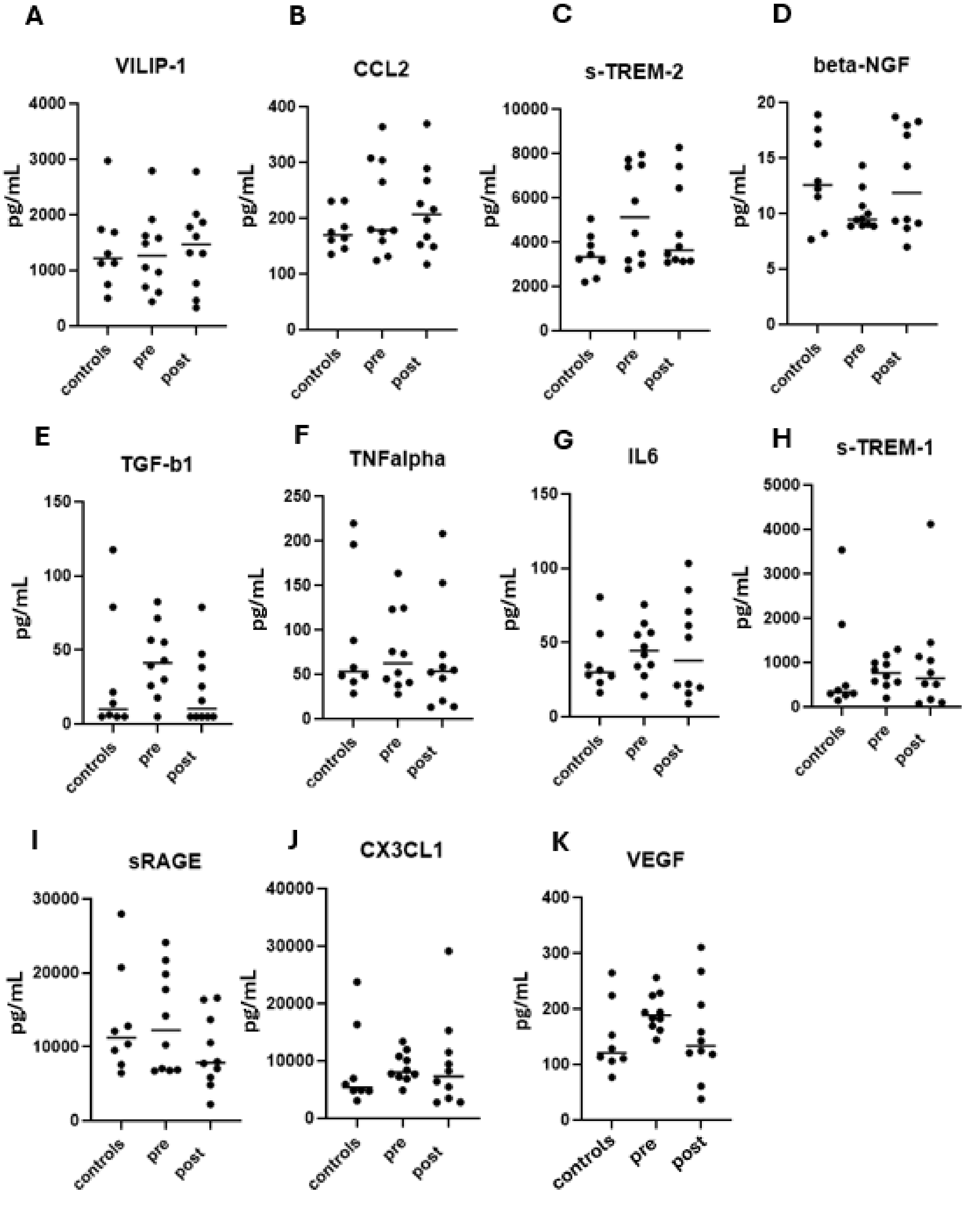


**Supplementary Figure 2. Plasma cytokine expression in PANS-pre and PANS-post compared to healthy controls.**

A-K: Plasma cytokine expression levels of healthy control children, PANS-pre, and PANS-post visinin-like protein 1 (VILIP-1), C-C motif ligand 2 (CCL2), soluble form of triggering receptor expressed on myeloid cells 2 (s-TREM-2), beta nerve growth factor (β-NGF), transforming growth factor beta (TGF-β1), tumour necrosis factor alpha (TNF-α), interleukin 6 (IL-6), s-TREM-1, soluble receptor for advanced glycation end products (sRAGE), chemokine (C-X3-C motif) ligand 1 (CX3CL1), and vascular endothelial growth factor (VEGF).

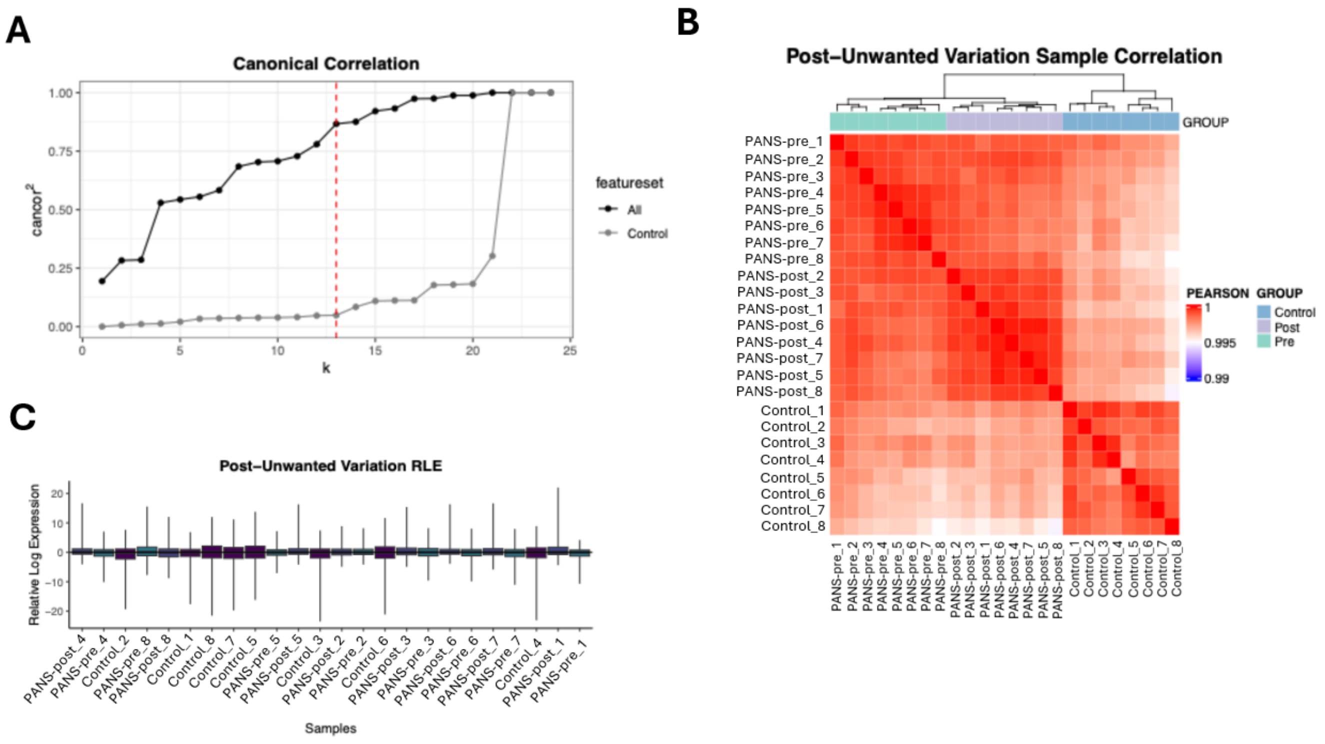


**Supplementary Figure 3. Exploratory bulk RNA sequencing and removal of unwanted variation.**

(A) In bulk RNA sequencing, k = 13 (factors of unwanted variation) were used to remove genes with minimal differential expression compared to negative control genes.

(B) Heatmap depicting high pearson correlation and clustering of samples within groups following removal of unwanted variation.

(C) Box and whisker relative log expression plot of samples after normalisation.

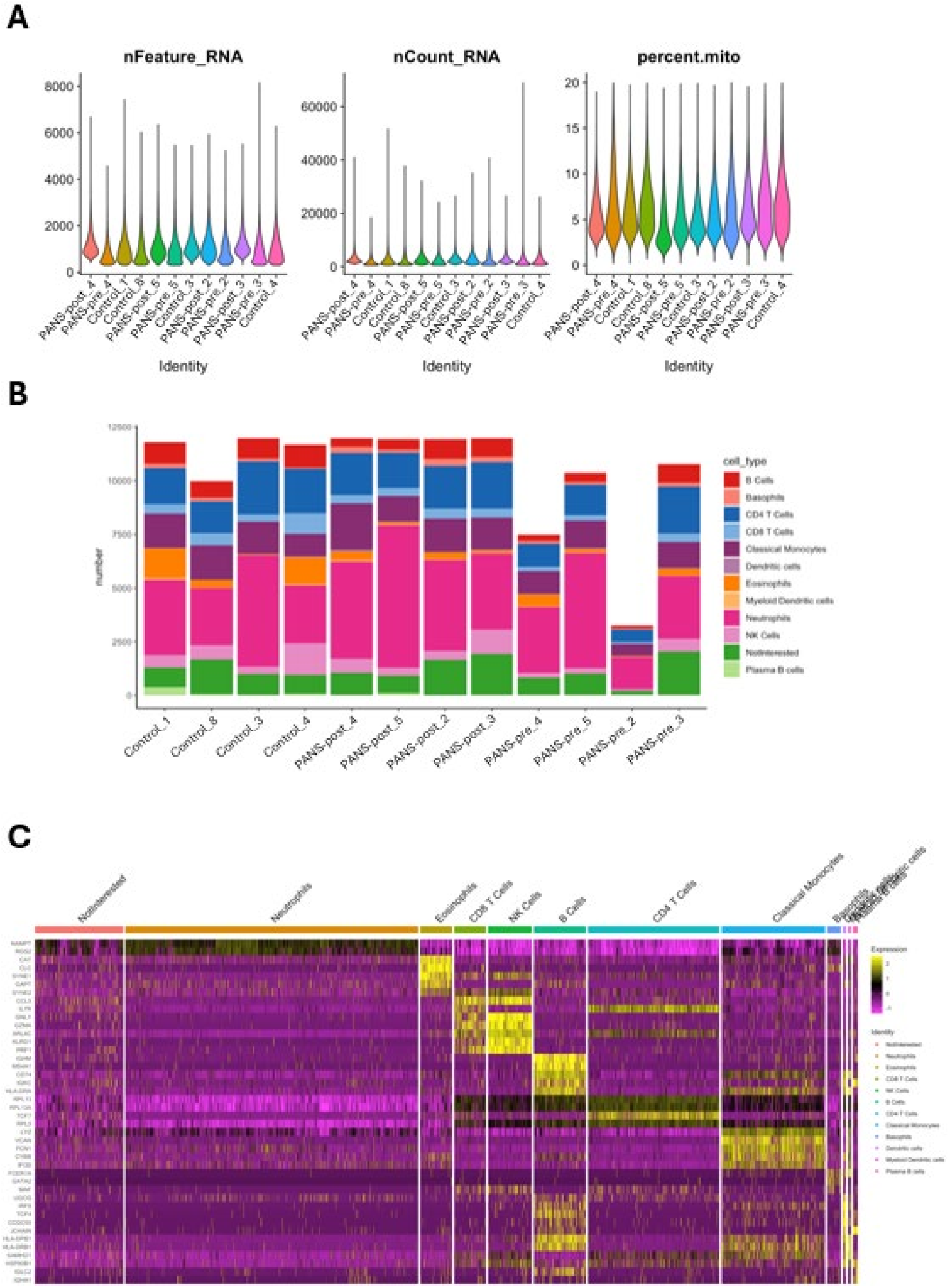


**Supplementary Figure 4. Single-cell RNA sequencing.**

(A) Number of features, counts, and percentage of mitochondrial genes per sample.

(B) Proportion of individual cell types per sample.

(C) Heat map of top differentially expressed genes based on cell type: genes are the top 5 markers of differential analysis between cell types (FDR <0.05) ranked by the difference in proportion of cells expressing the respective gene.

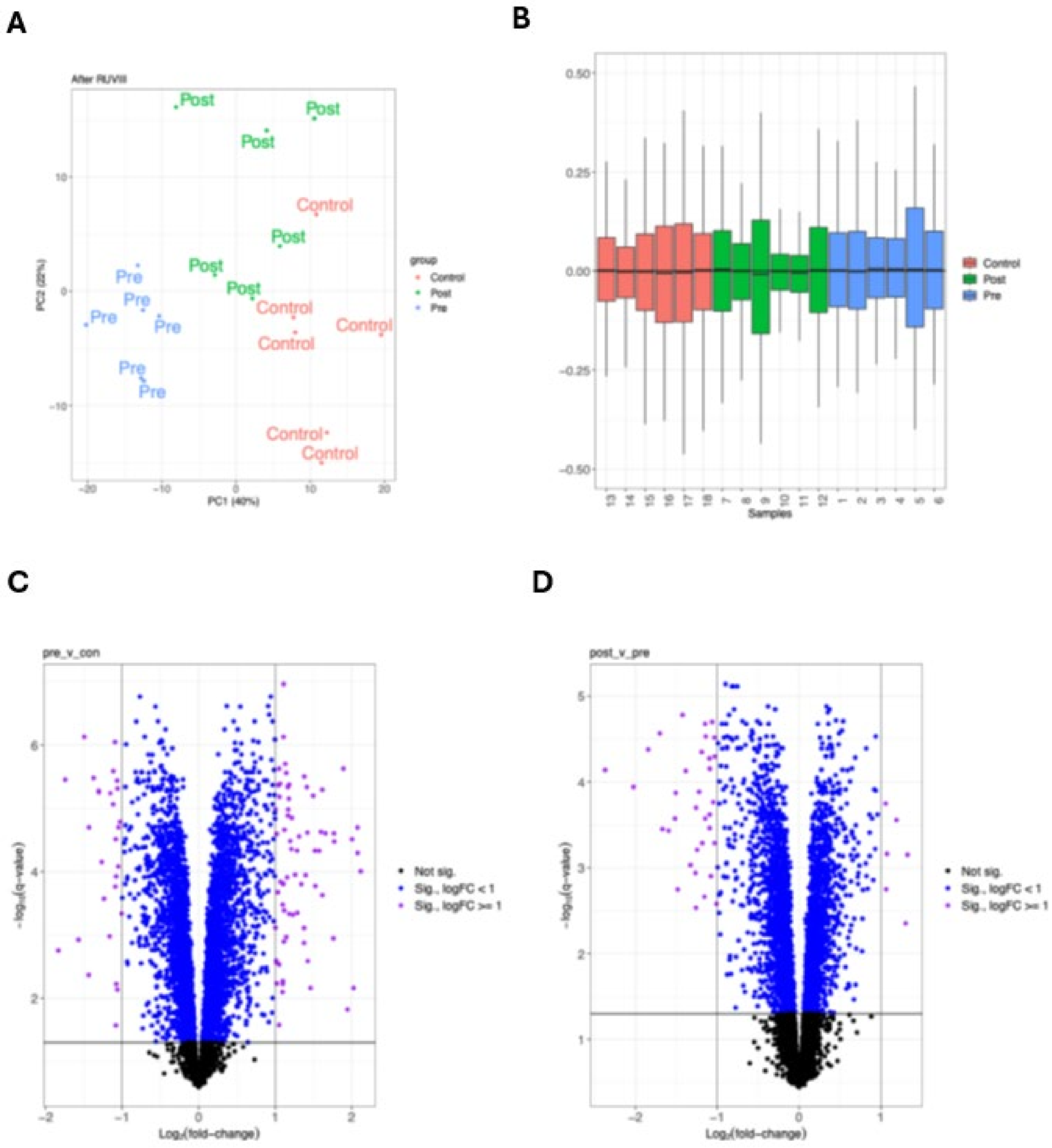


**Supplementary Figure 5. Proteomics analyses.**

(A) Principal component analysis (PCA) performed on proteomics shows clear discrimination between healthy controls, PANS-pre and PANS-post. The x-axis represents Principal Component 1 (PC1), while the y-axis represents Principal Component 2 (PC2).

(B) Box and whiskers relative log expression (y-axis) plot of samples after normalisation.

(C) Volcano plot of differentially expressed proteins (FDR <0.05) in the PANS-pre vs control comparison.

(D) Volcano plot of differentially expressed proteins (FDR <0.05) in the PANS-post vs pre comparison.

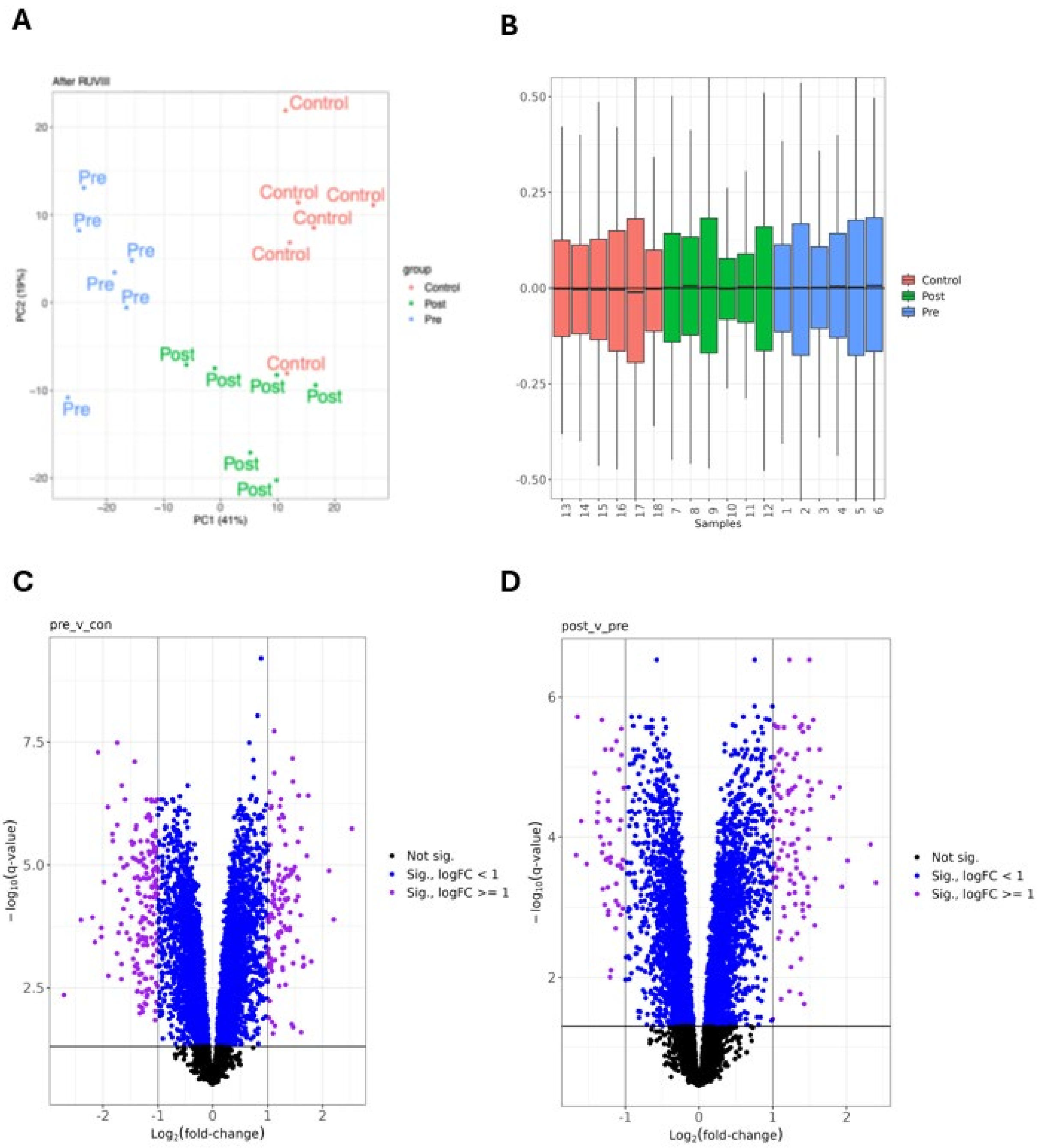


**Supplementary Figure 6. Phosphoproteomics analyses.**

(A) Principal component analysis (PCA) performed on phosphoproteomics shows clear discrimination between healthy controls, PANS-pre and PANS-post. The x-axis represents Principal Component 1 (PC1), while the y-axis represents Principal Component 2 (PC2).

(B) Box and whiskers relative log expression plot (y-axis) of samples after normalisation.

(C) Volcano plot of differentially expressed phosphopeptides (FDR <0.05) in the PANS-pre vs control comparison.

(D) Volcano plot of differentially expressed phosphopeptides (FDR <0.05) in the PANS-post vs pre comparison.

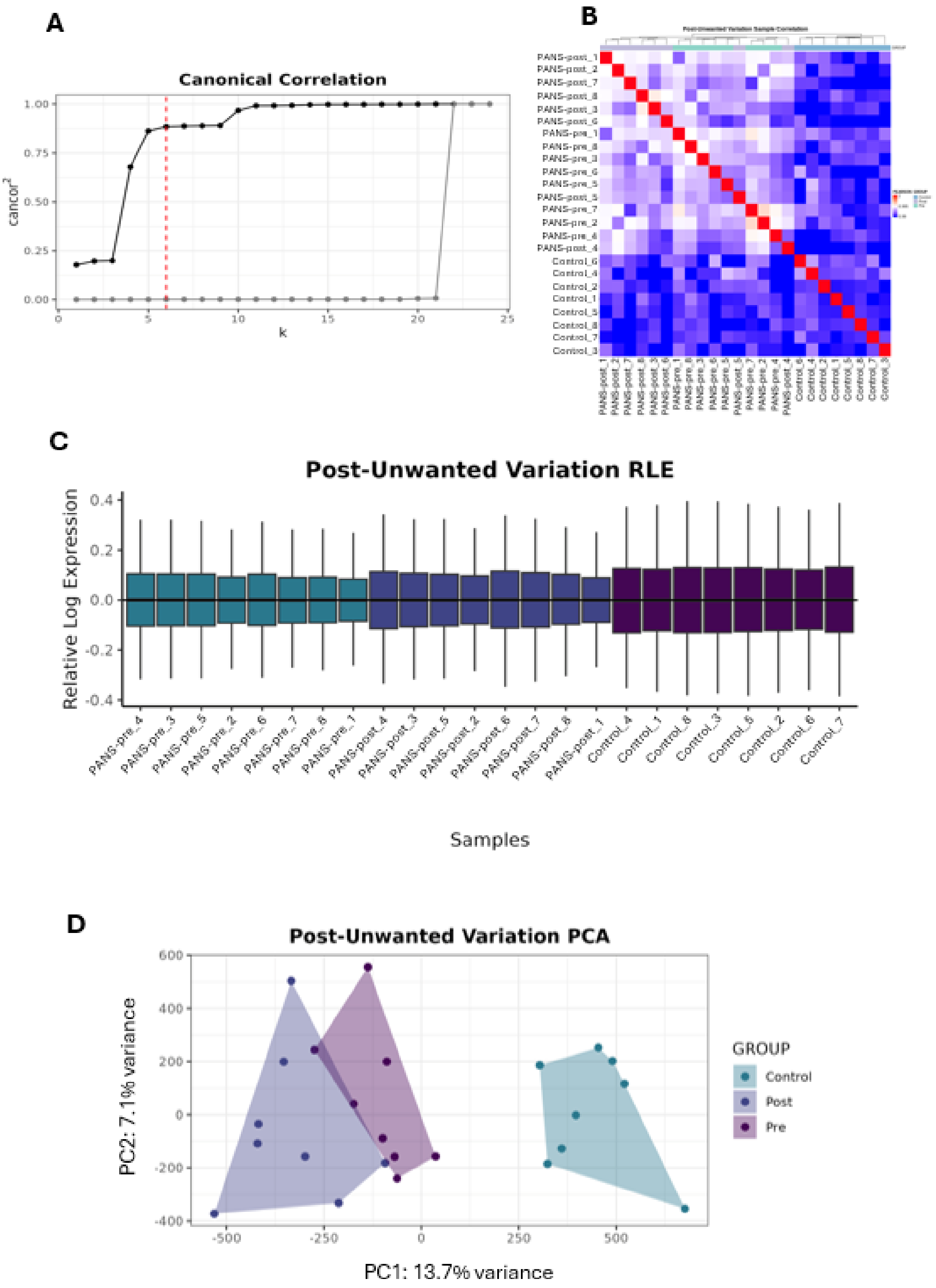


**Supplementary Figure 7. Methylation analyses.**

(A) For methylation data, k = 6 (factors of unwanted variation) were used to remove methylated positions with minimal differential expression compared to negative control methylated positions.

(B) Heatmap depicting high pearson correlation and clustering of samples within groups following removal of unwanted variation.

(C) Box and whisker relative log expression plot of samples after normalisation.

(D) Principal component analysis (PCA) performed on DNA methylation data shows clear discrimination between healthy controls, PANS-pre and PANS-post. The x-axis represents Principal Component 1 (PC1), while the y-axis represents Principal Component 2 (PC2).
